# An overview of current mental health in the general population of Australia during the COVID-19 pandemic: Results from the COLLATE project

**DOI:** 10.1101/2020.07.16.20155887

**Authors:** Susan L Rossell, Erica Neill, Andrea Phillipou, Eric J Tan, Wei Lin Toh, Tamsyn E Van Rheenen, Denny Meyer

## Abstract

The novel coronavirus disease (COVID-19) poses significant mental health challenges globally; however, to date, there is limited community level data. This study reports on the first wave of data from the COLLATE project (COvid-19 and you: mentaL heaLth in AusTralia now survEy), an ongoing study aimed at understanding the impact of the COVID-19 pandemic on the mental health and well-being of Australians. This paper addresses prevailing primary concerns related to the COVID-19 pandemic, current levels of negative emotions and risk factors predicting these negative emotions. On April 1st to 4th 2020, 5158 adult members of the Australian general public completed an online survey. Participants ranked their top ten current primary concerns about COVID-19, and completed standardized measures to ascertain levels of negative emotions (specifically, depression, anxiety and stress). Socio-demographic information was also collected and used in the assessment of risk factors. The top three primary concerns were all related to the health and well-being of family and loved ones. As expected, levels of negative emotion were exceptionally high. Modelling of predictors of negative emotions established several risk factors related to demographic variables, personal vulnerabilities, financial stresses, and social distancing experiences; particularly being young, being female, or having a mental illness diagnosis. The data provides important characterization of the current mental health of Australians during the COVID-19 pandemic. Critically, it appears that specific groups in the Australian community may need special attention to ensure their mental health is protected during these difficult times. The data further suggests the need for immediate action to combat high levels of psychological distress, along with the exacerbation of mental health conditions, in relation to the COVID-19 pandemic in Australia. These results may provide some direction for international researchers hoping to characterize similar issues in other countries.

## Introduction

The novel coronavirus disease (COVID-19) emerged in China in late 2019 and has spread rapidly across the globe. It is a contagious viral infection presenting with respiratory, neurological, gastrointestinal, and cardiac symptoms that range in severity from non-symptomatic through to causing death (Mao et al., 2020). The first case in Australia reported symptoms on January 13th 2020 (2019-nCoV National Incident Room Surveillance Team, 2020). This was followed by an exponential increase in infections, and unfortunately, deaths (the first Australian death occurred on February 24th). Since COVID-19 was declared a pandemic by the World Health Organization (WHO) on March 11th 2020 (WHO, 2020), the world has been engulfed in an unprecedented global crisis characterized by threatened or actual healthcare system collapse, job losses, and a failing global economy. In Australia, the crisis has been compounded by the implementation of government-regulated restrictions to contain the virus affecting social liberties.

In light of this, COVID-19 poses a significant mental health challenge to the Australian population, both now and in the long term. However, at the time of conducting this project there was no community level data in relation to the mental health implications of the COVID-19 pandemic in Australia, and only one published study from another Western country. In that study of 1310 adults in Spain, heightened negative emotions were associated with being female, being younger, and having negative self-perceptions (Losada-Baltar et al., 2020). Four studies on the psychological impacts of COVID-19 have also emerged from China. One study analyzed Weibo (Chinese social media platform) posts from 17,865 active users using online ecological recognition based on machine-learning predictive models (Li et al., 2020). The results showed that negative emotions increased (e.g., anxiety, depression and indignation), while positive emotions (e.g., happiness) and life satisfaction decreased over a two-week period from January 13th to January 26th 2020. Two other studies compared the psychological status of medical and non-medical (administration) health workers, illustrating increased insomnia, fear, anxiety, depression, somatization, and obsessive-compulsive symptoms in medical health workers (Lu et al., 2020, Zhang et al., 2020). Living in rural areas, being female, and being in contact with COVID-19-positive patients were reported as risk factors for negative emotions. Finally, Wang et al. (Wang et al., 2020), using an online survey and snowball sampling in the general population between January 31st and February 2nd 2020, reported that ∼50% of the 1210 respondents rated the psychological impact of COVID-19 as moderate-to-severe, with 33% reporting moderate-to-severe anxiety. Student status, being female and poor self-rated health were reported as risk factors for negative emotions.

In Australia, several health and economic measures had been implemented by March 31st 2020, in an attempt to control the spread of COVID-19 and stave off economic recession (e.g. $130b towards keeping Australians employed). While a recent funding announcement of $1.1b to boost digital mental health services is welcome, if we are to adequately manage this COVID-19 mental health crisis, there is a time-critical need to empirically characterize the current psychological impacts of the pandemic on the Australian population. This is particularly relevant given the Australian Government’s current implementation of ‘social distancing’, a key transmission-prevention measure that describes the maintenance of minimum physical space between oneself and those outside of one’s home. Social distancing restrictions, which limit one’s out-of-home movements unless absolutely essential, have been found to increase social isolation and loneliness (Zhang et al., 2020), alcohol abuse (Wu et al., 2008), and domestic violence (Galea et al., 2020). This could translate to widespread fear, anxiety, and depression in general society, particularly exacerbated in persons with existing mental health conditions who have an increased susceptibility to the adverse impacts of stress (Duan and Zhu, 2020).

Here we report on the first wave of data collected from the COLLATE project (COvid-19 and you: mentaL heaLth in AusTralia now survEy), an ongoing study aimed at understanding the impact of the COVID-19 pandemic on the mental health and wellbeing of Australians. The COLLATE project (described below) focuses on identifying the current concerns, emotional experiences and risk factors for adverse COVID-19-related mental health outcomes in people currently living in Australia. In our initial analysis of wave 1 data, we focused on characterizing the primary concerns of respondents related to the current COVID-19 pandemic as of April 1st to 4th 2020. Levels of negative emotion (depression, anxiety, and stress) were examined and compared with existing Australian population norms; and were modelled as an outcome to identify possible risks factors related to demographic variables, personal vulnerabilities, financial stresses, and social distancing experiences.

## Methods

This study received ethics approval from Swinburne University Human Ethics Review Committee (approval number: 20202917-4107) and complied with the Declaration of Helsinki.

### Study Design and Population

On April 1st 2020, adult members of the Australian general public (aged 18+) were invited to participate in an anonymous ∼15-20mins online survey, completed at their convenience (i.e. the inclusion criteria to participate were being aged 18+ years and currently residing in Australia). Participants were informed that 16 surveys would be issued over the course of the project. These would be active for 72 hours per month, from 9am on the 1st to 8:59am on the 4th (Australian Eastern Standard Time), occurring monthly for the first year and then annually for the subsequent four years (Tan et al., 2020). Participants were informed that they could complete as many or as few surveys as they wanted, with surveys from the same respondent being linked by a personalized pseudonym (thus a subsample would provide us with longitudinal data, with the remaining data cross-sectional snapshots over the 16 surveys).

After online consent, participants completed the survey which covered three broad topics: a) current concerns, b) current emotional experiences, and c) socio-demographics/risk factors. Items from previously validated surveys were incorporated where possible, in line with good practice in survey creation (Thayer-Hart, Dykema, Elver, Schaeffer, & Stevenson, 2010). Relevant existing scales and measures were included if they had good reliability/validity. Demographic items were included based on examinations of other large-scale Australian surveys (including the Household, Income and Labor Dynamics in Australia (HILDA) Survey, the National Drug Strategy Household Survey and the Domestic and Family Violence Survey, the Australian Bureau of Statistics National Health Survey) additional items were created where necessary to ensure that all areas of interest were covered. In terms of item structure, many of the demographic questions were multiple choice or check box options. For more exploratory items, open ended questions with text boxes for responses were provided. As noted, the data described here relate to survey round 1: April 2020, and only the measures addressing our aims for this manuscript are described below.

Invitations to complete the survey were placed on digital university and community noticeboards and social media (e.g. Facebook, LinkedIn, Instagram, and Twitter) as well as participant registries held within the Centre for Mental Health at Swinburne University, which included participants with identified mental health conditions. Exponential non-discriminative snowball sampling was used, with all participants asked to pass the invitation onto their networks.

### Measures

#### Primary concerns

Participants were asked to identify and rank their top 10 current concerns (out of 23) relating to the COVID-19 pandemic, with 1 being their greatest concern (see Table 2 for the full list of concerns).

#### Negative emotions

The Depression Anxiety Stress Scale (DASS-21) was used. It is a 21-item self-report measure yielding three subscales – depression, anxiety, and stress – each containing seven items (Lovibond and Lovibond, 1995). Individual items are scored on a four-point Likert scale (0 to 3). DASS-21 raw scores were doubled to render them comparable to full-length DASS scores (42 items).

#### Risk factors

Measurement of risk factors were divided into four categories: *Demographics* including: age, gender, education, living situation, geographical location/state, whether born in Australia, ethnicity, and religion; *Personal vulnerabilities* including: being someone at increased mortality risk (e.g. immune-compromised, >60years), having lived experience of mental illness, being a carer of someone with a mental illness or special needs, and being a healthcare professional or ‘essential’ worker; *Financial stresses*: fortnightly take-home pay, cash savings, mortgage repayments/rent, self-employment, job loss, and occupation; *Social distancing experiences*: perceived positives of the situation, perception of government restrictions on mental health, perception of social distancing measures duration, and working from home.

### Statistical Analyses

Data were analyzed in *SPSS v26*.*0*. For all analyses, weights were used to adjust for imbalances in the sample based on the Australian Bureau of Statistics (ABS) population data for age, gender and geographical location/state (ABS, 2016). In all, there were 12 categories for age (18-19; 20-24; 25-29; 30-34; 35-39; 40-44; 45-49; 50-54; 55-59; 60-64; 65-70; 70+), two categories for gender (male; female) and four categories for state (Victoria; New South Wales; Queensland; Australian Capital Territory + Northern Territory + Western Australia + South Australia + Tasmania). By March 31st 2020 23:59 Australian Eastern Standard Time (AEST) there were 4,707 confirmed cases of COVID-19 in Australia, with 18 deaths. The majority of confirmed cases were in the states of New South Wales (n=2,182), Queensland (n=743) and Victoria (n=917), with the other states and territories reporting a total of 865 cases together. Given confirmed case numbers, we stratified by state by examining these three states independently from the other states and territories, which were combined.

#### Primary concerns

To characterize the top ten primary concerns, the number of respondents endorsing each concern was obtained and mean rankings were computed for the ten most commonly selected options. Rankings of zero were assigned to options not endorsed by a participant, and rankings of 1 to 10 were computed for endorsed concerns, with 10 for the option of greatest concern. In this case, the weights developed used the joint distributions of the three weighting variables ensuring that the sum of weights assigned was 5545 (i.e. the total number of respondents to this question who also provided age, gender and geographical location/state data).

#### Negative emotions

Depression, anxiety, and stress subscales and total DASS scores were compared to Australian population norms (Lovibond and Lovibond, 1995) using *t*-tests. Additionally, the percentage of participants (weighted and non-weighted) were calculated across the four defined severity levels (normal, mild, moderate, severe/extremely severe) for the three negative emotions. Respondents who failed to complete more than 10% of the DASS items were removed from the analysis. Remaining missing items were imputed using the EM algorithm as Little’s MCAR test showed items were missing completely at random. For this analysis, the sum of weights and sample size were equal to 5158.

#### Risk factors

Using a transformed (SQRT) total DASS, the relationships between negative emotions and the four domains (*demographics, personal vulnerabilities, financial stresses* and *social distancing experiences*) were explored using general linear model analyses.

## Results

### Sample description

8014 participants started the survey, with *n*=5545 respondents (∼30% attrition) completing the primary concerns ranking question and providing demographic data. For the negative emotion analyses *n*=5158 respondents completed the DASS. Demographic data is displayed in Table 1. The sample was biased in favour of females aged 25-45 living in the state of Victoria, making the use of post-stratification weighting essential in subsequent analyses.

**Table 1:**
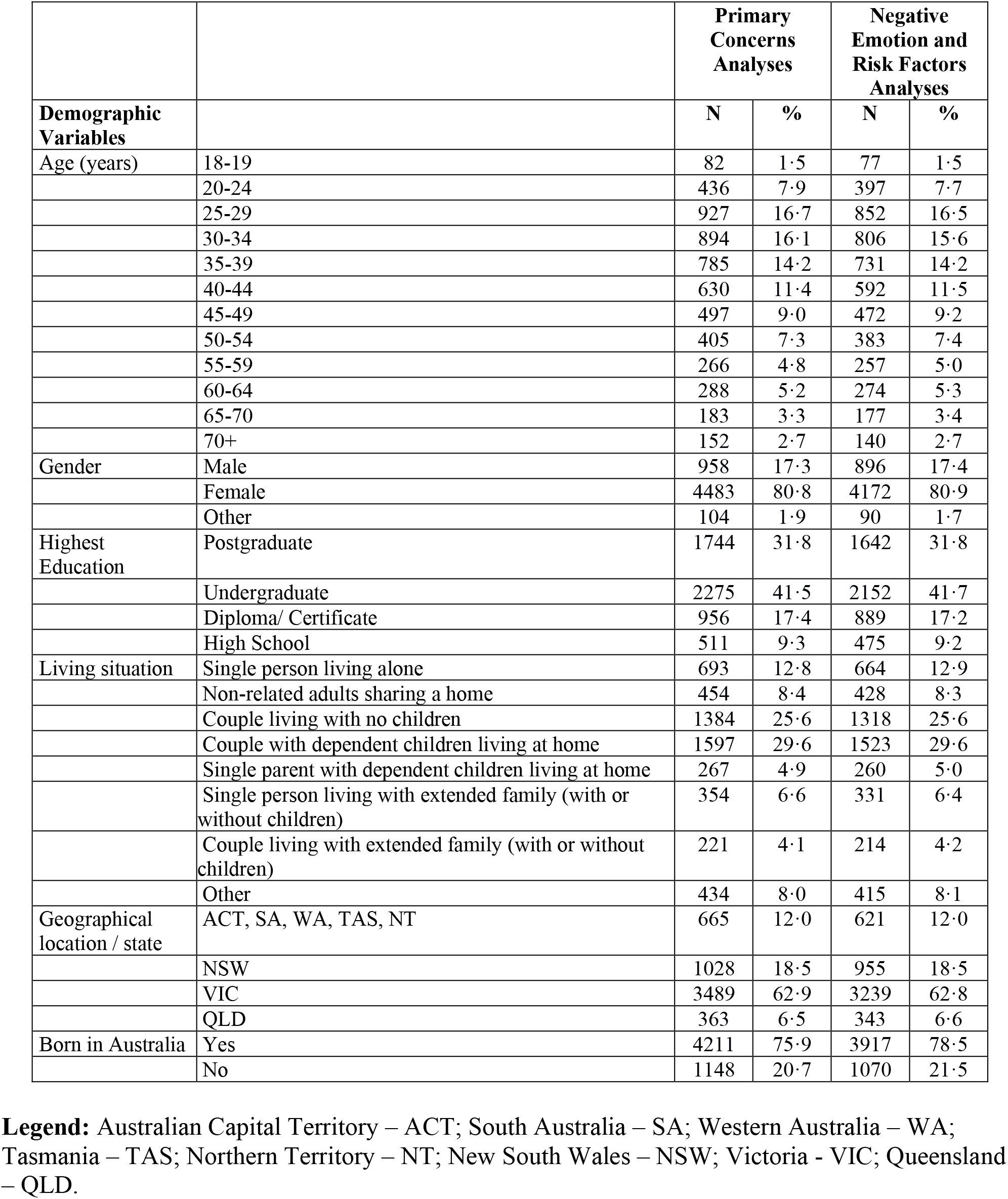
Sociodemographic description of the sample.

|  |  | Primary Concerns Analyses |  | Negative Emotion and Risk Factors Analyses |  |
| --- | --- | --- | --- | --- | --- |
|  |  | N | % | N | % |
| **Demographic Variables** |  |  |  |  |  |
| Age (years) | 18-19 | 82 | 1.5 | 77 | 1.5 |
|  | 20-24 | 436 | 7.9 | 397 | 7.7 |
|  | 25-29 | 927 | 16.7 | 852 | 16.5 |
|  | 30-34 | 894 | 16.1 | 806 | 15.6 |
|  | 35-39 | 785 | 14.2 | 731 | 14.2 |
|  | 40-44 | 630 | 11.4 | 592 | 11.5 |
|  | 45-49 | 497 | 9.0 | 472 | 9.2 |
|  | 50-54 | 405 | 7.3 | 383 | 7.4 |
|  | 55-59 | 266 | 4.8 | 257 | 5.0 |
|  | 60-64 | 288 | 5.2 | 274 | 5.3 |
|  | 65-70 | 183 | 3.3 | 177 | 3.4 |
|  | 70+ | 152 | 2.7 | 140 | 2.7 |
| Gender | Male | 958 | 17.3 | 896 | 17.4 |
|  | Female | 4483 | 80.8 | 4172 | 80.9 |
|  | Other | 104 | 1.9 | 90 | 1.7 |
| Highest Education | Postgraduate | 1744 | 31.8 | 1642 | 31.8 |
|  | Undergraduate | 2275 | 41.5 | 2152 | 41.7 |
|  | Diploma/ Certificate | 956 | 17.4 | 889 | 17.2 |
|  | High School | 511 | 9.3 | 475 | 9.2 |
| Living situation | Single person living alone | 693 | 12.8 | 664 | 12.9 |
|  | Non-related adults sharing a home | 454 | 8.4 | 428 | 8.3 |
|  | Couple living with no children | 1384 | 25.6 | 1318 | 25.6 |
|  | Couple with dependent children living at home | 1597 | 29.6 | 1523 | 29.6 |
|  | Single parent with dependent children living at home | 267 | 4.9 | 260 | 5.0 |
|  | Single person living with extended family (with or without children) | 354 | 6.6 | 331 | 6.4 |
|  | Couple living with extended family (with or without children) | 221 | 4.1 | 214 | 4.2 |
|  | Other | 434 | 8.0 | 415 | 8.1 |
| Geographical location / state | ACT, SA, WA, TAS, NT | 665 | 12.0 | 621 | 12.0 |
|  | NSW | 1028 | 18.5 | 955 | 18.5 |
|  | VIC | 3489 | 62.9 | 3239 | 62.8 |
|  | QLD | 363 | 6.5 | 343 | 6.6 |
| Born in Australia | Yes | 4211 | 75.9 | 3917 | 78.5 |
|  | No | 1148 | 20.7 | 1070 | 21.5 |

**Legend:** Australian Capital Territory – ACT; South Australia – SA; Western Australia – WA; Tasmania – TAS; Northern Territory – NT; New South Wales – NSW; Victoria - VIC; Queensland – QLD.

### Primary concerns

The primary concern data, with the percentage of respondents rating their top 10 concerns, is presented in Table 2. Mean rankings were ordered from 1 to 23 in declining order of importance (mean and standard deviations calculated for the rankings of each concern).

“Implications for health and wellbeing of family loved ones” was the most commonly endorsed concern, however, it ranked 3rd in terms of mean rankings. “Loved one dying of coronavirus” and “Loved one catching coronavirus” were the next most commonly endorsed primary concerns, and had the highest mean rankings.

**Table 2:**
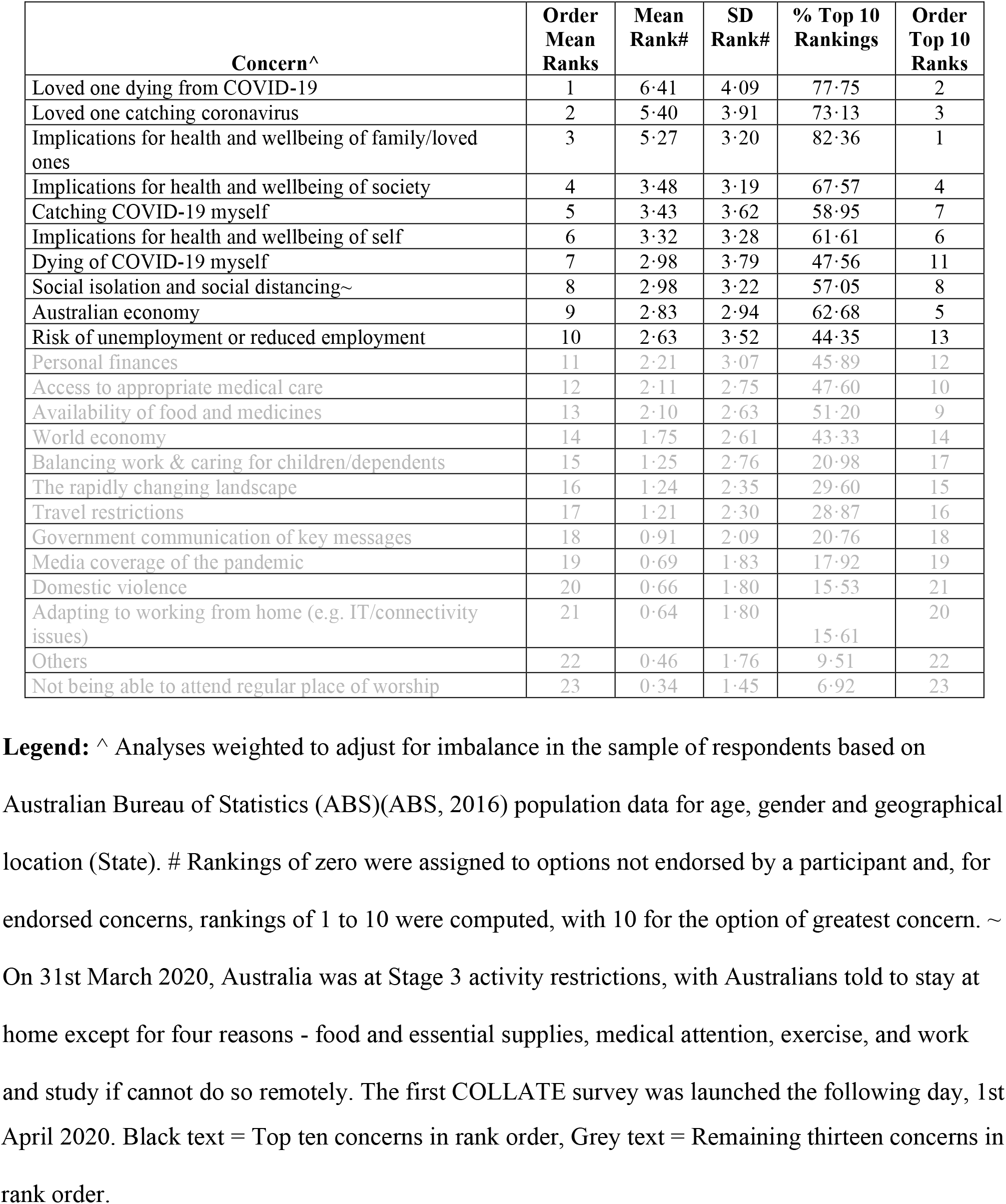
Rank data illustrating the ten primary concerns endorsed by Australians (N= 5545)

| Concern^ | Order Mean Ranks | Mean Rank# | SD Rank# | % Top 10 Rankings | Order Top 10 Ranks |
| --- | --- | --- | --- | --- | --- |
| Loved one dying from COVID-19 | 1 | 6.41 | 4.09 | 77.75 | 2 |
| Loved one catching coronavirus | 2 | 5.40 | 3.91 | 73.13 | 3 |
| Implications for health and wellbeing of family/loved ones | 3 | 5.27 | 3.20 | 82.36 | 1 |
| Implications for health and wellbeing of society | 4 | 3.48 | 3.19 | 67.57 | 4 |
| Catching COVID-19 myself | 5 | 3.43 | 3.62 | 58.95 | 7 |
| Implications for health and wellbeing of self | 6 | 3.32 | 3.28 | 61.61 | 6 |
| Dying of COVID-19 myself | 7 | 2.98 | 3.79 | 47.56 | 11 |
| Social isolation and social distancing~ | 8 | 2.98 | 3.22 | 57.05 | 8 |
| Australian economy | 9 | 2.83 | 2.94 | 62.68 | 5 |
| Risk of unemployment or reduced employment | 10 | 2.63 | 3.52 | 44.35 | 13 |
| Personal finances | 11 | 2.21 | 3.07 | 45.89 | 12 |
| Access to appropriate medical care | 12 | 2.11 | 2.75 | 47.60 | 10 |
| Availability of food and medicines | 13 | 2.10 | 2.63 | 51.20 | 9 |
| World economy | 14 | 1.75 | 2.61 | 43.33 | 14 |
| Balancing work & caring for children/dependents | 15 | 1.25 | 2.76 | 20.98 | 17 |
| The rapidly changing landscape | 16 | 1.24 | 2.35 | 29.60 | 15 |
| Travel restrictions | 17 | 1.21 | 2.30 | 28.87 | 16 |
| Government communication of key messages | 18 | 0.91 | 2.09 | 20.76 | 18 |
| Media coverage of the pandemic | 19 | 0.69 | 1.83 | 17.92 | 19 |
| Domestic violence | 20 | 0.66 | 1.80 | 15.53 | 21 |
| Adapting to working from home (e.g. IT/connectivity issues) | 21 | 0.64 | 1.80 | 15.61 | 20 |
| Others | 22 | 0.46 | 1.76 | 9.51 | 22 |
| Not being able to attend regular place of worship | 23 | 0.34 | 1.45 | 6.92 | 23 |

**Legend:** ^ Analyses weighted to adjust for imbalance in the sample of respondents based on Australian Bureau of Statistics (ABS)(ABS, 2016) population data for age, gender and geographical location (State). # Rankings of zero were assigned to options not endorsed by a participant and, for endorsed concerns, rankings of 1 to 10 were computed, with 10 for the option of greatest concern. ~ On 31st March 2020, Australia was at Stage 3 activity restrictions, with Australians told to stay at home except for four reasons - food and essential supplies, medical attention, exercise, and work and study if cannot do so remotely. The first COLLATE survey was launched the following day, 1st
April 2020. Black text = Top ten concerns in rank order, Grey text = Remaining thirteen concerns in rank order.

### Negative emotions

Figure 1 (also see Supplementary Table 1) shows mean values for the DASS scores compared to Australian norms. In all cases, they were significantly greater than the norms (*p*s<.001); with people self-identifying as having a mental health diagnosis (MH DX) scoring 5-5.5-fold higher than those without such a diagnosis, who themselves scored 3 times higher than normative levels. Table 3 presents the score distributions across the four severity levels (normal, mild, moderate, severe/extremely severe); 21-35% of the population demonstrated moderate-to-extremely severe depression, anxiety and stress.

**Table 3:** DASS scores across the severity levels in the current Australian data from COLLATE in comparison to the Chinese data from Wang et al. (Wang et al., 2020)

|  |  | Depression |  |  | Anxiety |  |  | Stress |  |  |
| --- | --- | --- | --- | --- | --- | --- | --- | --- | --- | --- |
|  |  | Aus W | Aus NW | China | Aus W | Aus NW | China | Aus W | Aus NW | China |
| Normal | N | 2796 | 2521 | 843 | 3480 | 3058 | 770 | 2328 | 1870 | 821 |
|  | % | 54.2% | 48.9% | 69.7% | 67.5% | 59.3% | 63.6% | 45.1% | 36.3% | 67.9% |
| Mild | N | 752 | 810 | 167 | 402 | 439 | 91 | 1730 | 1825 | 292 |
|  | % | 14.6% | 15.7% | 13.8% | 7.8% | 8.5% | 7.5% | 33.5% | 35.4% | 24.1% |
| Moderate | N | 922 | 1040 | 148 | 713 | 892 | 247 | 725 | 929 | 66 |
|  | % | 17.9% | 20.2% | 12.2% | 13.8% | 17.3% | 20.4% | 14.1% | 18.0% | 5.5% |
| Severe/Extremely Severe | N | 687 | 787 | 52 | 562 | 770 | 102 | 375 | 534 | 31 |
|  | % | 13.3% | 15.3% | 4.3% | 10.9% | 14.9% | 8.4% | 7.3% | 10.4% | 2.6% |

**Legend:** Aus W = Australia weighted data, Aus NW = Australia non-weighted data; Depression: normal (score: 0-6), mild (score: 10-12), moderate (score: 13-20), severe/extremely severe (score: 21-42); Anxiety: normal (score: 0-6), mild (score: 7-9), moderate (score: 10-14), severe/extremely severe (score: 15-42) and Stress: normal (score: 0-10), mild (score: 11-18), moderate (score: 19-26), severe/extremely severe (score: 27-42). Scores as per Lovibond & Lovibond (Lovibond and Lovibond, 1995), it should be noted that DASS is not a categorical measure of clinical diagnosis, for clinical purposes it can be helpful to have ‘labels’ to characterize degree of severity relative to the population.

**Figure 1:**
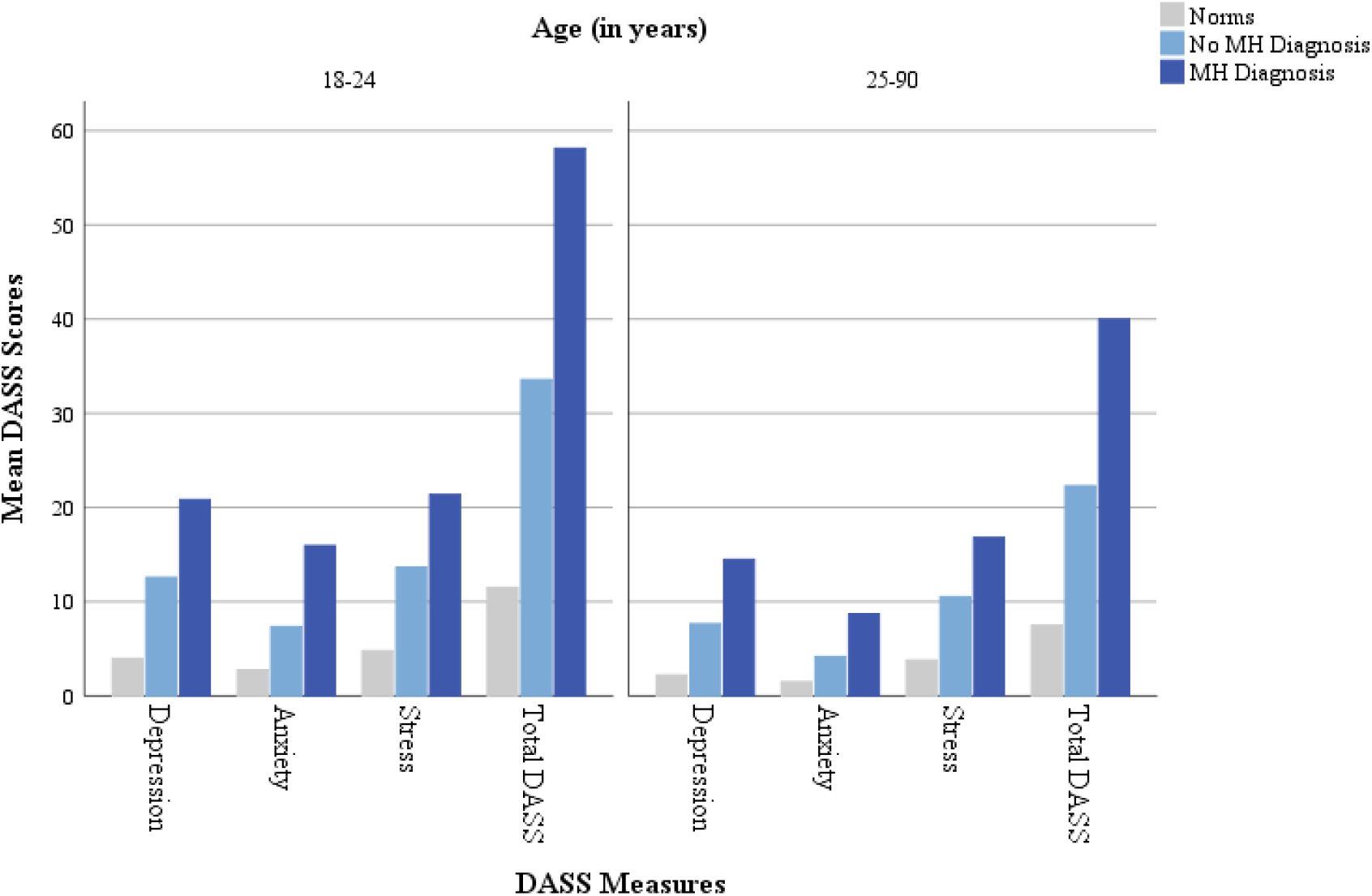
Comparison of weighted sample results with existing Australian norms for the DASS-21. **Legend:** Norms in Australia from Lovibond & Lovibond (Lovibond and Lovibond, 1995), MH = Mental Health

### Risk factors

#### Demographics

18.7% of the variation in negative emotions was explained by demographic factors (Supplementary Table 2). Lower levels of negative emotions were demonstrated by: people aged 30-34 and 70+, males, people with higher levels of education, couples (with or without children), and non-Australian born residents. The two states with the most COVID-19 cases (i.e. New South Wales and Victoria) showed lower negative emotions than the other states and territories.

#### Personal vulnerabilities

Adding personal vulnerabilities to the initial model explained an additional 10.5% of the variation in negative emotions (Supplementary Table 3). People with a higher mortality risk, lived experience of mental illness, carer responsibilities for someone with a mental illness or special needs, as well as people in “essential” occupations, all had higher levels of negative emotions.

#### Financial stresses

Adding finance-related variables to the model explained an additional 5.2% of the variation (Supplementary Table 4). Individuals with higher fortnightly incomes and cash savings demonstrated lower levels of negative emotions. Higher levels of negative emotions were experienced by people under financial stress to meet mortgage and rental payments as well as those expecting to lose their jobs. Additionally, highest negative emotions were present for the unemployed, closely followed by homemakers, volunteers, or retired people.

#### Social distancing experiences

An additional 8.4% of the variation was explained by adding social distancing variables (Supplementary Table 5). Generally speaking, higher negative emotions were recorded for those who found they now had more free time. This included those who had more down-time, more time to spend communicating with family, more time to do jobs around the house and for those who identified no positive influences in the current situation. However, negative emotions were lower for those who found they now had more time for hobbies. Negative emotions were higher for those who reported that the government restrictions were adversely impacting their mental health, and for those who thought that the current restrictions might continue for more than 12 months. Finally, negative emotions were higher for those not working from home.

#### Risk factor summary

The four domains explained 42.8% of the variance in negative emotions (summarized in Table 4). Important predictors for high negative emotions (i.e. η^2^≥0.010) were being young (18-24), being female, being single, living in states with lower COVID-19 cases (QLD, ACT, SA, WAS, TAS, NT), being at higher risk of mortality and having a lived experience of mental illness. The perceived negative effect of government restrictions on mental health was also highly associated with negative emotions, demonstrating the largest effect size, η^2^=0.102. Having sizeable cash savings, owning one’s own home and predicting a short duration of the current situation were protective factors against experiencing negative emotions.

**Table 4:** *R*-Square values and most important predictor variables for negative emotions.

| Predictor Domains | Increase in R-Square | R-Square | Most important predictors |
| --- | --- | --- | --- |
| Demographics | 18.7% | 18.7% | Age ($\eta^2=0.099$)<br>Gender ($\eta^2=0.027$)<br>Living situation ($\eta^2=0.011$)<br>State ($\eta^2=0.016$) |
| Personal vulnerabilities | 10.5% | 29.2% | Higher mortality risk ($\eta^2=0.020$)<br>Lived experience mental health illness ($\eta^2=0.095$) |
| Financial stresses | 5.2% | 34.4% | Home rental stress ($\eta^2=0.012$)<br>Level of cash savings ($\eta^2=0.012$) |
| Social distancing experiences | 8.2% | 42.8% | Effects of government restrictions ($\eta^2=0.102$)<br>Expected duration of current situation ($\eta^2=0.010$) |

**Legend:** All selected predictors with partial effect sizes $\eta^2 \geq 0.010$

## Discussion

The first wave of data from the COLLATE project provides an important characterization of the current mental health of Australians during the COVID-19 pandemic. The top three primary concerns among the general public were all related to the health and well-being of family and loved ones, specifically loved ones catching or dying from COVID-19. As expected, levels of negative emotions (depression, anxiety and stress) were exceptionally high, approximately three times greater than existing population norms in those with no pre-existing mental health conditions. Of concern was the finding that those with a pre-existing mental health condition demonstrated negative emotions 5 to 5.5 times greater than population norms. When the current Australian DASS data was compared with Chinese data (Wang et al., 2020), two differences emerged. First, the mean total DASS score from China of 20.16 (SD 20.42) was lower than that of Australia, even for individuals not reporting a mental health condition (18-24years: 33.56 (SD 25.49) and 25years+: 22.29 (SD 16.90)). Second, more Australians were classified as having moderate-to-extremely severe negative emotions (see Table 3). These apparent cross-cultural differences will need to be further investigated with a specifically designed comparison study, with differences in social norms and civil liberties between the two countries being possible influences. Nonetheless, both the Australian and Chinese data speak to the elevation of negative emotions in the general population during the COVID-19 pandemic.

In our data, modelling predictors of negative emotions established several risks factors related to demographic variables, personal vulnerabilities, financial stresses, and social distancing experiences. This included young people (18-24 years) as well as those that are approaching middle-age (35-50 years). Given that the number of young people experiencing mental health conditions has been rising over the last decade in Australia (Carlisle et al., 2019), and internationally (Miron et al., 2019), this current data of such high levels of negative emotions in young people (up to age 24 here) is of particular concern. Increased negative emotions in our middle-aged respondents were associated with increased childcare duties and/or financial stresses that are specific to the immediate situation. This speaks to the importance of monitoring negative emotions in both young people and the middle-aged group longitudinally, that is, in the short and long term.

Being female was another significant risk factor for high levels of negative emotions (Lu et al., 2020, Wang et al., 2020, Zhang et al., 2020). While possible reasons for this remain to be determined, we speculate this could relate to juggling work and increased childcare duties, heightened risk of being in a domestic violence situation, as well as the higher risks of job loss and/or higher likelihood of being an ‘essential’ worker. Those under financial strain and those who were unemployed are also at increased risk of psychological distress. Thus, methods for targeting this ‘financial strain’ population to offer them more affordable options for mental health support will be important.

Finally, those with pre-existing mental illness are of specific concern (Phillipou et al., 2020; Neill et al, 2020; Van Rheenen et al, 2020), and existing mental health services will likely need increased support to meet the rising needs of these consumers. It is notable that since this survey was conducted, the Australian government has announced a significant boost in funding to support mental health ($76m over the next two years) with initiatives including dedicated websites and phone lines to support people experiencing stress and anxiety from prevailing COVID-19 related pressures, as well as a public information campaign. The current findings strongly underlie the need for such initiatives to be more targeted to specific groups.

Two other findings in our data warrant discussion. First, respondents from the two states with the highest number of COVID-19 cases, New South Wales and Victoria, were found to have lower negative emotions. While this was unexpected, further examination of risk factors established there were a number of protective personal vulnerabilities and financial stresses for persons living in these two states. That is, these states encompassed a lower percentage of respondents with lived experience of a mental illness and a greater percentage of respondents with financial stability. Another important, albeit unsurprising, finding from our data was that individuals who perceived that current government restrictions were very negatively impacting their mental health also had the most pronounced negative emotions. In the context of respondents’ primary concerns with the health and well-being of family members, this finding reinforces that government restrictions, such as social distancing, may be better be framed in public messaging as necessary for protecting loved ones and ourselves from contracting the virus. Such a refocusing on positive outcomes may provide individuals with a sense of agency that tempers the powerlessness of being given a legal mandate to stay home.

A limitation of the study was the snowballing approach to survey recruitment; this resulted in a non-representative sample of the Australian population, which included some respondents with known mental health diagnosis. To address this, weightings were used based on ABS data (ABS, 2016) to statistically correct for any bias. However, even with statistical weighting, it is difficult to account for specific subgroups, for example those without access to the internet. Furthermore, despite >8000 participants starting the survey, only ∼5500 had useable datasets due to considerable attrition (30%), which is albeit typical of online research. This data provides a snapshot of mental health and well-being of Australians in April 2020 in relation to COVID-19; to do so we compared current negative emotions to existing Australian norms. It is possible differences in sampling factors related to the current sample data and existing norms may explain some of the differences rather than COVID-19 itself. However, given the magnitude of our findings in terms of elevated negative emotions in the general community such sampling differences are unlikely to explain the large variance between current and norm data. We also did not assess the mental health of school-aged children and adolescents in the current survey, the majority are whom are being home-schooled and not permitted face-to-face contact with their peers in Australia at present. School-age children may thus be particularly vulnerable to stress and anxiety, and a critical avenue for future work will be to examine this group specifically.

## Conclusion

The data collected from the COLLATE project will provide a reference for healthcare professionals in terms of current mental health needs in Australia, in addition to guiding policymakers in making accurate provisions within mental health services and actionable policies. The findings are predicted to be applicable across other nations with similar healthcare systems and government management of the COVID-19 pandemic. Important findings from the data are that: a) people with existing mental health conditions have very high levels of negative emotions (as per (Duan and Zhu, 2020)), in addition, there are b) high levels of psychological distress in the general community, with some individuals particularly vulnerable (as per (Wang et al., 2020)). The fluid nature of the situation throughout this pandemic makes our continuing and longitudinal comparisons of the mental health effects of government restrictions and lock-down duration forecasts a priority for future study. Overall, this data has made it clear that increased mental health support will be of paramount importance as the world faces the consequences of the COVID-19 pandemic.

## Data Availability

The data analysis scripts and results files used for this paper are available for review on request by qualified researchers-scientists. All requests for future use of the dataset require a concept proposal describing the purpose of data access, appropriate ethical approval, and provision for data security

## Acknowledgements

SLR holds a Senior National Health and Medical Research Council (NHMRC) Fellowship (GNT1154651), and EJT (GNT1142424) and TVR (GNT1088785) hold Early Career NHMRC Fellowships. AP (GNT1159953) and WLT (GNT1161609) are supported by NHMRC New Investigator Project Grants. The authors would also like to thank all the participants who took the time to participate in this study.

## Author Contributions

SLR conceived the project and the design. All authors SLR, DM, EN, AP, EJT, WLT and TVR finalized the design, constructed the survey, obtained ethics, engaged in data collection and interpretation of findings. DM completed all the data analyses in consultation with the other authors. All authors prepared the manuscript and agreed to its final form.

## Supplementary Material

**Supplementary Table 1:**
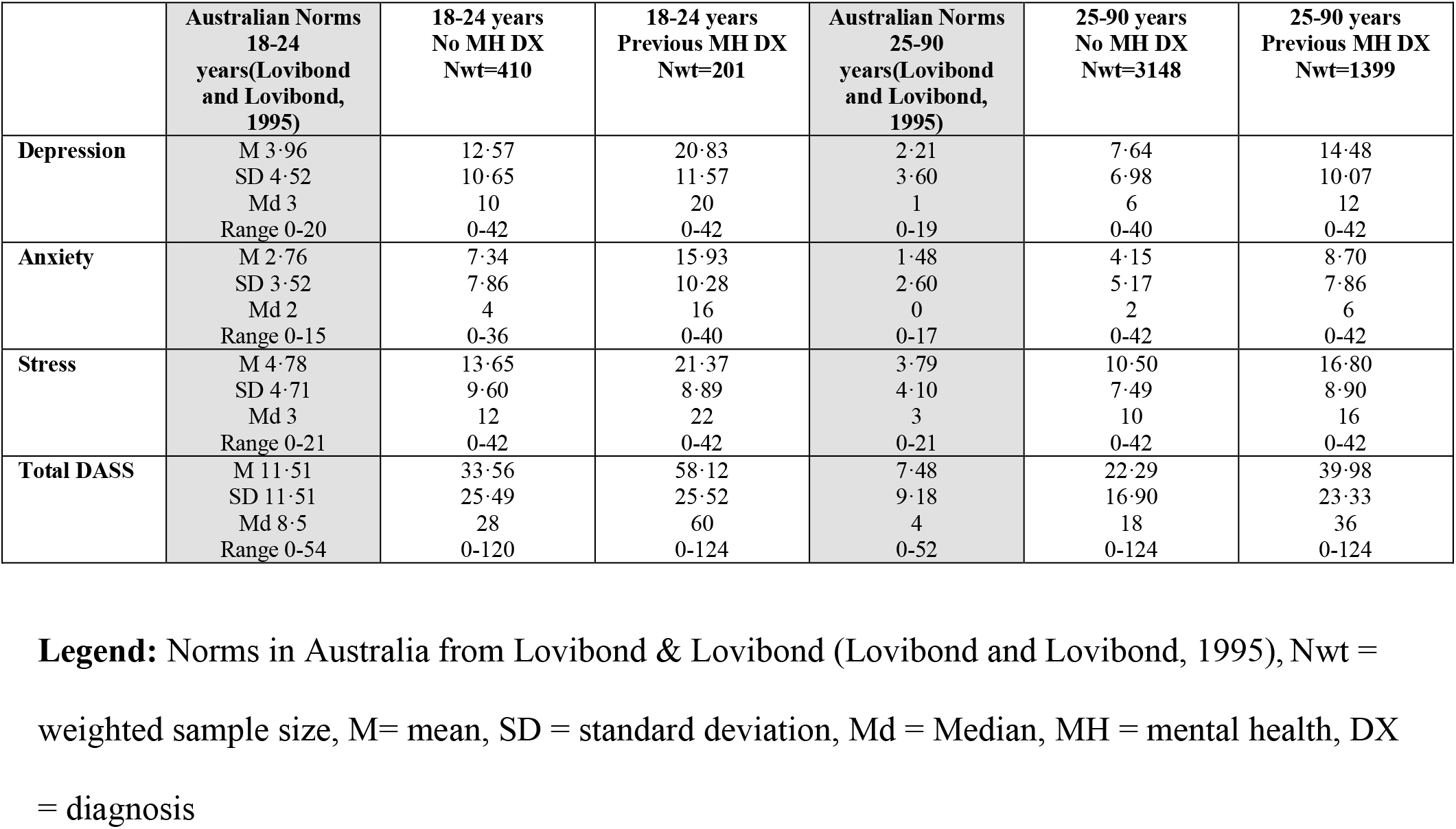
Comprehensive comparison of weighted sample results with existing Australian norms for the DASS-21.

|  | Australian Norms<br>18-24<br>years(Lovibond<br>and Lovibond,<br>1995) | 18-24 years<br>No MH DX<br>Nwt=410 | 18-24 years<br>Previous MH DX<br>Nwt=201 | Australian Norms<br>25-90<br>years(Lovibond<br>and Lovibond,<br>1995) | 25-90 years<br>No MH DX<br>Nwt=3148 | 25-90 years<br>Previous MH DX<br>Nwt=1399 |
| --- | --- | --- | --- | --- | --- | --- |
| **Depression** | M 3.96<br>SD 4.52<br>Md 3<br>Range 0-20 | 12.57<br>10.65<br>10<br>0-42 | 20.83<br>11.57<br>20<br>0-42 | 2.21<br>3.60<br>1<br>0-19 | 7.64<br>6.98<br>6<br>0-40 | 14.48<br>10.07<br>12<br>0-42 |
| **Anxiety** | M 2.76<br>SD 3.52<br>Md 2<br>Range 0-15 | 7.34<br>7.86<br>4<br>0-36 | 15.93<br>10.28<br>16<br>0-40 | 1.48<br>2.60<br>0<br>0-17 | 4.15<br>5.17<br>2<br>0-42 | 8.70<br>7.86<br>6<br>0-42 |
| **Stress** | M 4.78<br>SD 4.71<br>Md 3<br>Range 0-21 | 13.65<br>9.60<br>12<br>0-42 | 21.37<br>8.89<br>22<br>0-42 | 3.79<br>4.10<br>3<br>0-21 | 10.50<br>7.49<br>10<br>0-42 | 16.80<br>8.90<br>16<br>0-42 |
| **Total DASS** | M 11.51<br>SD 11.51<br>Md 8.5<br>Range 0-54 | 33.56<br>25.49<br>28<br>0-120 | 58.12<br>25.52<br>60<br>0-124 | 7.48<br>9.18<br>4<br>0-52 | 22.29<br>16.90<br>18<br>0-124 | 39.98<br>23.33<br>36<br>0-124 |

**Legend:** Norms in Australia from Lovibond & Lovibond (Lovibond and Lovibond, 1995), Nwt = weighted sample size, M= mean, SD = standard deviation, Md = Median, MH = mental health, DX = diagnosis

